# Iron deficiency testing among people with incident heart failure in primary care

**DOI:** 10.64898/2026.06.14.26355616

**Authors:** Vijay Maharajan, Nicholas R Jones, Clare Bankhead, Innocent Erone, Sarah Haynes, Alvin M Katumba, Christien Ka Hou Li, Suzanne Maynard, Noemi BA Roy, Akshay Shah, Simon J Stanworth, Margaret Smith, Cynthia Wright Drakesmith

## Abstract

**Background:** Given around 50% of people with heart failure have a degree of iron deficiency, guidelines recommend screening. It is uncertain to what extent this is done in primary care and whether testing is equitable.

**Aim:** To report the proportion of people with incident heart failure who undergo a ferritin test within 12 months.

**Design and setting:** Retrospective primary care cohort study using Clinical Practice Research Datalink Aurum data, between 2016 and 2021.

**Methods:** We report the proportion of adults with an incident diagnosis of heart failure who received a ferritin test within 12 months. Multivariable logistic regression was used to examine the odds of testing based on key demographic covariates and co-morbidities.

**Results:** Among 105,749 individuals with an incident diagnosis of heart failure (mean age 71.6 years, SD 14.3), only 35,688 (33.7%) received a ferritin test within the subsequent year. Increasing age (odds ratio 1.25 per 10-year increase, 95% CI: 1.24–1.27), female sex (male sex OR 0.86, 0.84–0.89) and Asian ethnicity (OR 1.70, 1.59–1.80) were all associated with increased odds of testing as were diagnoses of coeliac disease (OR 1.86, 1.58-2.21), type 1 diabetes (OR 1.82, 1.51-2.19) and cirrhosis (OR 1.64, 1.43-1.87). There was geographic variation in testing, even in adjusted analyses.

**Conclusion:** In a large primary care dataset, two thirds of people with incident heart failure did not receive a ferritin test for iron deficiency within a year of diagnosis demonstrating a gap in current practice and an opportunity for improvements in service delivery.

## Introduction

Around 50% of patients with chronic heart failure have varying degrees of iron deficiency, which may be caused by decreased absorption of iron from the gut due to oedema, increased loss, or impaired iron metabolism linked to chronic inflammation.^1^ ^2^ Iron deficiency, irrespective of the presence of anaemia, has important clinical consequences; it is associated with increased disease severity including elevated natriuretic peptides levels and New York Heart Association class.^3^ Iron deficiency is an independent predictor of both all-cause and cardiovascular mortality in heart failure and is also associated with reduced exercise capacity and worse quality of life.^4^ ^5^

Randomised trials have reported that treatment with intravenous ferric carboxymaltose for iron deficiency and heart failure with reduced ejection fraction (HFrEF) can improve functional capacity and quality of life.^6–8^ A meta-analysis of 3,773 patients with heart failure and iron deficiency across ten randomised trials reported that intravenous iron replacement also resulted in a reduction in the composite primary outcome of cardiovascular death or hospitalisation for heart failure (hazard ratio 0.75, 95% confidence interval 0.61 to 0.93).^9^ The National Institute for Health and Care Excellence (NICE) and European Society of Cardiology (ESC) both now recommend considering intravenous iron replacement for people with HFrEF who have iron deficiency, defined as a serum ferritin <100 ng/mL or a serum ferritin between 100-299 ng/mL if TSAT is below 20%, independent of the presence of anaemia.^10–12^

Clinicians are therefore recommend to screen for iron deficiency in patients with heart failure to inform both the prognostic assessment and treatment,^11^ ^13^ with European guidance recommending repeat testing once or twice per year.^14^ The extent to which these guidelines are being implemented in UK primary care is unknown. This is important when considering equitable access to treatment and opportunities for improvement in service delivery. The primary aim of this study was to report the proportion of patients with an incident diagnosis of heart failure recorded in their primary care record who had a serum ferritin checked within the subsequent year. We also report the proportion of people who had a repeat ferritin test within a year.

## Methods

### Study design and setting

We conducted a retrospective primary care cohort study using Clinical Practice Research Datalink (CPRD) Aurum with permission from the CPRD’s Expert Review Committee (eRAP number 22_001873). The NHS Research Ethics Committee provides ethical approval to allow researchers to use anonymised patient data through CPRD. CPRD Aurum includes over 1,800 English GP practices and over 40 million patient records and is broadly representative of the wider UK population.^15^ We present these results in accordance with The REporting of studies Conducted using Observational Routinely-collected health Data (RECORD) Statement (Supplementary material).^16^

### Participants

Eligible patients were aged ≥18 years with an incident diagnostic code for heart failure recorded in their primary care electronic healthcare record (EHR) between January 2016 and March 2021. The cohort start date was chosen to coincide with the publication of the 2016 ESC Heart Failure Guidelines, which first recommend testing for iron deficiency.^13^ We only included participants with at least 12 months of follow-up data to exclude individuals with a very poor prognosis or receiving palliative care, where iron testing and treatment may not be appropriate.

### Variables

The primary outcome was a coded ferritin value within the primary care record within the first year following an incident heart failure diagnosis. Implausible ferritin values, defined as negative or >100,000 µg/L, were excluded. Demographic data were collected including age, sex, ethnicity, Index of Multiple Deprivation (IMD) and geographic region within England. Ethnicity was reported into five categories based on the Census 2021 approach.^17^ IMD was categorised into quintiles from 1 (least deprived) to 5 (most deprived). CPRD’s practice-level geographic variable was used to categorise general practices by regions.

We considered that patterns of ferritin testing may be different among patients with other long-term health conditions. In particular, we were interested in chronic conditions with an inflammatory component, as these can both complicate the interpretation of ferritin and influence the likelihood of iron testing and treatment. To identify these, we extracted the top 1,000 most frequently recorded Medcode IDs in CPRD, then manually grouped these into clinically relevant categories. These groups were reviewed by a team of clinicians to focus on conditions with plausible links to iron metabolism or historically established pathways for iron testing, resulting in a final list of 19 chronic inflammatory or systemic conditions (see Table 1). In combination these were considered multiple long-term conditions (MLTCs). Standardised codelists were created for each condition and independently reviewed by two clinicians. Patients were flagged as having a condition if a diagnosis was recorded prior to their incident heart failure date.

**Table 1.**
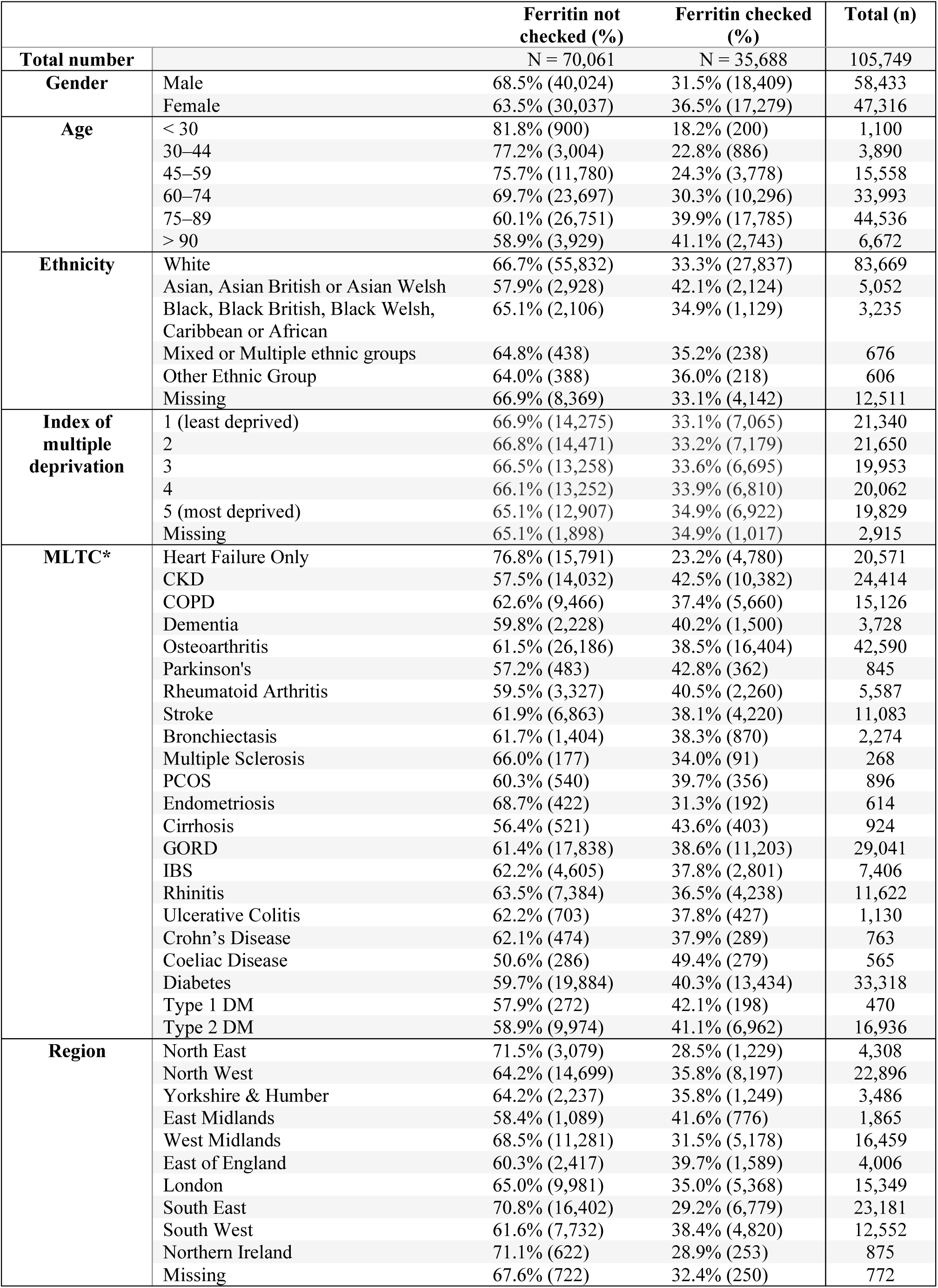
Baseline characteristics of patients with incident heart failure, categorised by whether subsequent ferritin testing was performed.

|  |  | **Ferritin not checked (%)** | **Ferritin checked (%)** | **Total (n)** |
| --- | --- | --- | --- | --- |
| **Total number** |  | N = 70,061 | N = 35,688 | 105,749 |
| **Gender** | Male | 68.5% (40,024) | 31.5% (18,409) | 58,433 |
|  | Female | 63.5% (30,037) | 36.5% (17,279) | 47,316 |
| **Age** | < 30 | 81.8% (900) | 18.2% (200) | 1,100 |
|  | 30–44 | 77.2% (3,004) | 22.8% (886) | 3,890 |
|  | 45–59 | 75.7% (11,780) | 24.3% (3,778) | 15,558 |
|  | 60–74 | 69.7% (23,697) | 30.3% (10,296) | 33,993 |
|  | 75–89 | 60.1% (26,751) | 39.9% (17,785) | 44,536 |
|  | > 90 | 58.9% (3,929) | 41.1% (2,743) | 6,672 |
| **Ethnicity** | White | 66.7% (55,832) | 33.3% (27,837) | 83,669 |
|  | Asian, Asian British or Asian Welsh | 57.9% (2,928) | 42.1% (2,124) | 5,052 |
|  | Black, Black British, Black Welsh, Caribbean or African | 65.1% (2,106) | 34.9% (1,129) | 3,235 |
|  | Mixed or Multiple ethnic groups | 64.8% (438) | 35.2% (238) | 676 |
|  | Other Ethnic Group | 64.0% (388) | 36.0% (218) | 606 |
|  | Missing | 66.9% (8,369) | 33.1% (4,142) | 12,511 |
| **Index of multiple deprivation** | 1 (least deprived) | 66.9% (14,275) | 33.1% (7,065) | 21,340 |
|  | 2 | 66.8% (14,471) | 33.2% (7,179) | 21,650 |
|  | 3 | 66.5% (13,258) | 33.6% (6,695) | 19,953 |
|  | 4 | 66.1% (13,252) | 33.9% (6,810) | 20,062 |
|  | 5 (most deprived) | 65.1% (12,907) | 34.9% (6,922) | 19,829 |
|  | Missing | 65.1% (1,898) | 34.9% (1,017) | 2,915 |
| **MLTC\*** | Heart Failure Only | 76.8% (15,791) | 23.2% (4,780) | 20,571 |
|  | CKD | 57.5% (14,032) | 42.5% (10,382) | 24,414 |
|  | COPD | 62.6% (9,466) | 37.4% (5,660) | 15,126 |
|  | Dementia | 59.8% (2,228) | 40.2% (1,500) | 3,728 |
|  | Osteoarthritis | 61.5% (26,186) | 38.5% (16,404) | 42,590 |
|  | Parkinson's | 57.2% (483) | 42.8% (362) | 845 |
|  | Rheumatoid Arthritis | 59.5% (3,327) | 40.5% (2,260) | 5,587 |
|  | Stroke | 61.9% (6,863) | 38.1% (4,220) | 11,083 |
|  | Bronchiectasis | 61.7% (1,404) | 38.3% (870) | 2,274 |
|  | Multiple Sclerosis | 66.0% (177) | 34.0% (91) | 268 |
|  | PCOS | 60.3% (540) | 39.7% (356) | 896 |
|  | Endometriosis | 68.7% (422) | 31.3% (192) | 614 |
|  | Cirrhosis | 56.4% (521) | 43.6% (403) | 924 |
|  | GORD | 61.4% (17,838) | 38.6% (11,203) | 29,041 |
|  | IBS | 62.2% (4,605) | 37.8% (2,801) | 7,406 |
|  | Rhinitis | 63.5% (7,384) | 36.5% (4,238) | 11,622 |
|  | Ulcerative Colitis | 62.2% (703) | 37.8% (427) | 1,130 |
|  | Crohn's Disease | 62.1% (474) | 37.9% (289) | 763 |
|  | Coeliac Disease | 50.6% (286) | 49.4% (279) | 565 |
|  | Diabetes | 59.7% (19,884) | 40.3% (13,434) | 33,318 |
|  | Type 1 DM | 57.9% (272) | 42.1% (198) | 470 |
|  | Type 2 DM | 58.9% (9,974) | 41.1% (6,962) | 16,936 |
| **Region** | North East | 71.5% (3,079) | 28.5% (1,229) | 4,308 |
|  | North West | 64.2% (14,699) | 35.8% (8,197) | 22,896 |
|  | Yorkshire & Humber | 64.2% (2,237) | 35.8% (1,249) | 3,486 |
|  | East Midlands | 58.4% (1,089) | 41.6% (776) | 1,865 |
|  | West Midlands | 68.5% (11,281) | 31.5% (5,178) | 16,459 |
|  | East of England | 60.3% (2,417) | 39.7% (1,589) | 4,006 |
|  | London | 65.0% (9,981) | 35.0% (5,368) | 15,349 |
|  | South East | 70.8% (16,402) | 29.2% (6,779) | 23,181 |
|  | South West | 61.6% (7,732) | 38.4% (4,820) | 12,552 |
|  | Northern Ireland | 71.1% (622) | 28.9% (253) | 875 |
|  | Missing | 67.6% (722) | 32.4% (250) | 772 |

### Statistical analysis

Descriptive statistics were used to describe the population, stratified by whether patients received a ferritin test within one year of heart failure diagnosis. Categorical variables including gender, ethnicity, IMD, region, and presence of MLTCs were summarised as counts and percentages.

Continuous variables were described using means and standard deviation or medians with interquartile ranges as appropriate. Age was categorised into 15-year bands to allow visual assessment of distribution. Multivariable logistic regression was then used to examine the associations of patient characteristics with the odds of receiving a ferritin test, adjusted for key demographic covariates. We repeated the regression analysis including each of the long-term conditions alone to examine the independent contribution to ferritin testing. Results are reported as odds ratios (ORs) or regression coefficients with 95% confidence intervals (CIs). A CONSORT-style flow diagram was constructed to illustrate patient inclusion and attrition across each stage of the analysis.

Among patients who had a baseline ferritin test, logistic regression was used to examine the associations of patient characteristics with the odds of receiving a repeat ferritin test within one year. Adjustment was made for age, sex, ethnicity, and region to allow comparability with the primary analysis. To estimate the adjusted effect of specific comorbidities, each MLTC was entered into a separate model with the same covariate adjustment.

Missing data were confined to ethnicity and geographic region, with the latter affecting only approximately 1,000 patients, suggesting that a small number of practices did not supply complete regional identifiers. Both variables were thought to be missing not at random, meaning multiple imputation was not considered appropriate and instead ‘missing’ was included as a separate category in regression models.^18^

All statistical analyses were conducted in Stata 18, following data extraction in SQL and pre-processing in Python.

## Results

Among over 47 million people within the database, we identified 105,749 with an incident diagnosis of heart failure within the study period (Figure 1). Mean age at diagnosis with heart failure was 71.6 years (SD 14.3) and 47,316 were female (44.7%) (Table 1). Patients tended to be older, with 48.4% (n=51,208) aged ≥75 years at the time of diagnosis compared to 19.4% (n=20,548) aged <60 years. The majority were of white ethnicity (n=83,669, 79.1%%). More people were diagnosed with incident heart failure from the least deprived quintile (n=21,340, 20.2%) compared to the most deprived (n=19,829, 18.8%). The most common of our pre-specified co-morbidities at the time of diagnosis included osteoarthritis (n=42,590, 40.3%), diabetes (n=33,318, 31.5%), gastro-oesophageal reflux disease (GORD) (n=29,041, 27.5%) and chronic kidney disease (CKD) (n=24,414, 23.1%).

**Figure 1.**
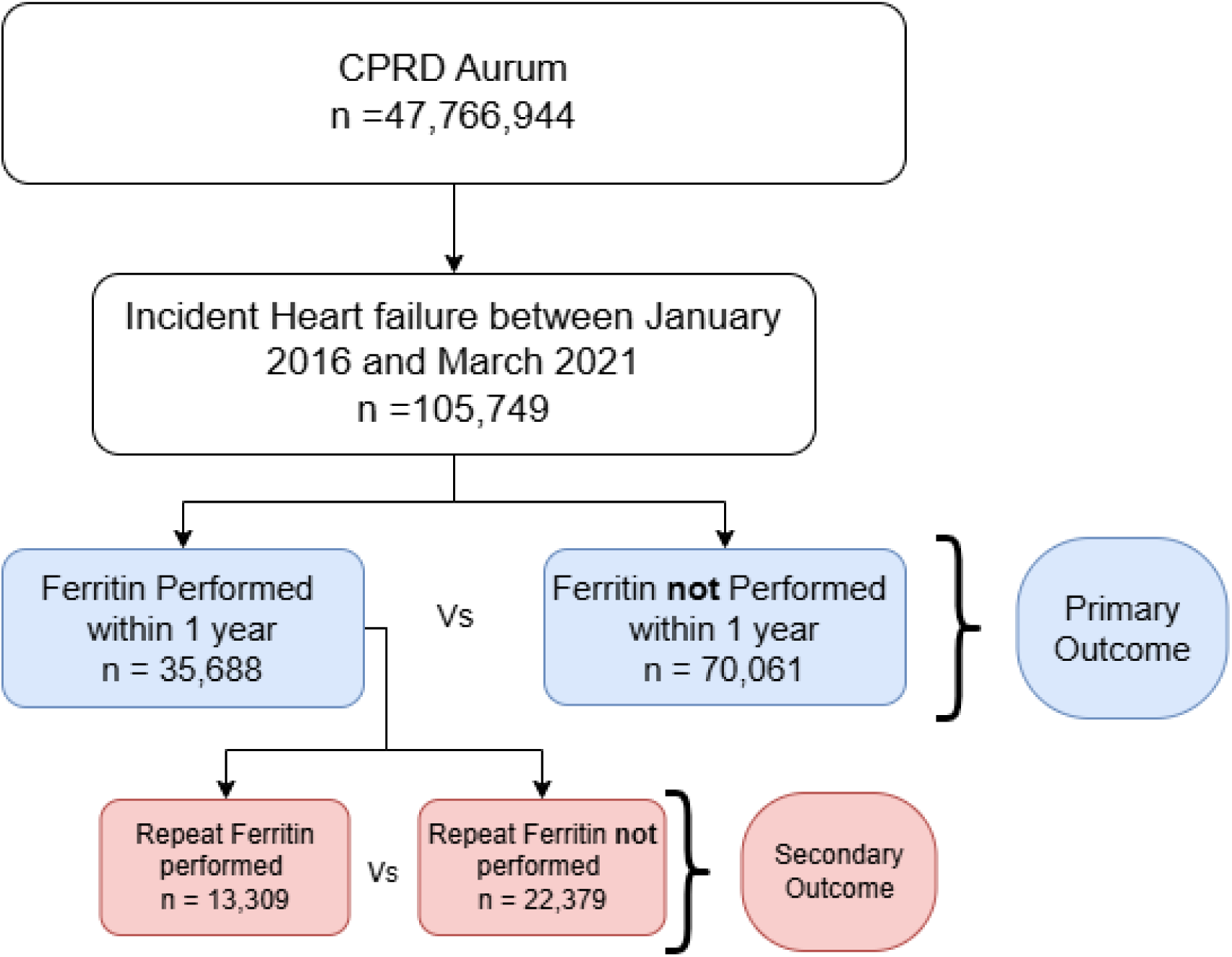
Flow diagram of study cohort.

### Patient factors associated with ferritin testing

Overall, 35,688 people received a ferritin test within a year of their heart failure diagnosis (33.7%). The proportion of people receiving a ferritin test increased with age and was highest in those aged ≥90 years (41.1%), followed by those aged 75 to 89 years (39.9%) and lowest in those aged under 30 years (18.2%) (Table 1). Almost half those who underwent a ferritin a test were aged 75–89 years (Figure 2). A slightly higher proportion of females received a ferritin test (36.5%) compared to males (31.5%) and a higher proportion of people of Asian ethnicity were tested (42.1%) than other ethnic groups, with the proportion of people tested lowest among white people (33.3%). A higher proportion of people in the most deprived IMD quintile received a test (n=6,922, 34.9%) than the least deprived (n=7,065, 33.1%). In regression analysis, there was higher odds of ferritin testing with increasing age (OR 1.25 per 10-year increase, 95% CI: 1.24–1.27), among people with Asian compared to White ethnicity (OR 1.70, 95% CI: 1.59–1.80) and lower odds of testing with male sex (OR 0.86, 95% CI: 0.84–0.89) (Table 2).

**Figure 2.**
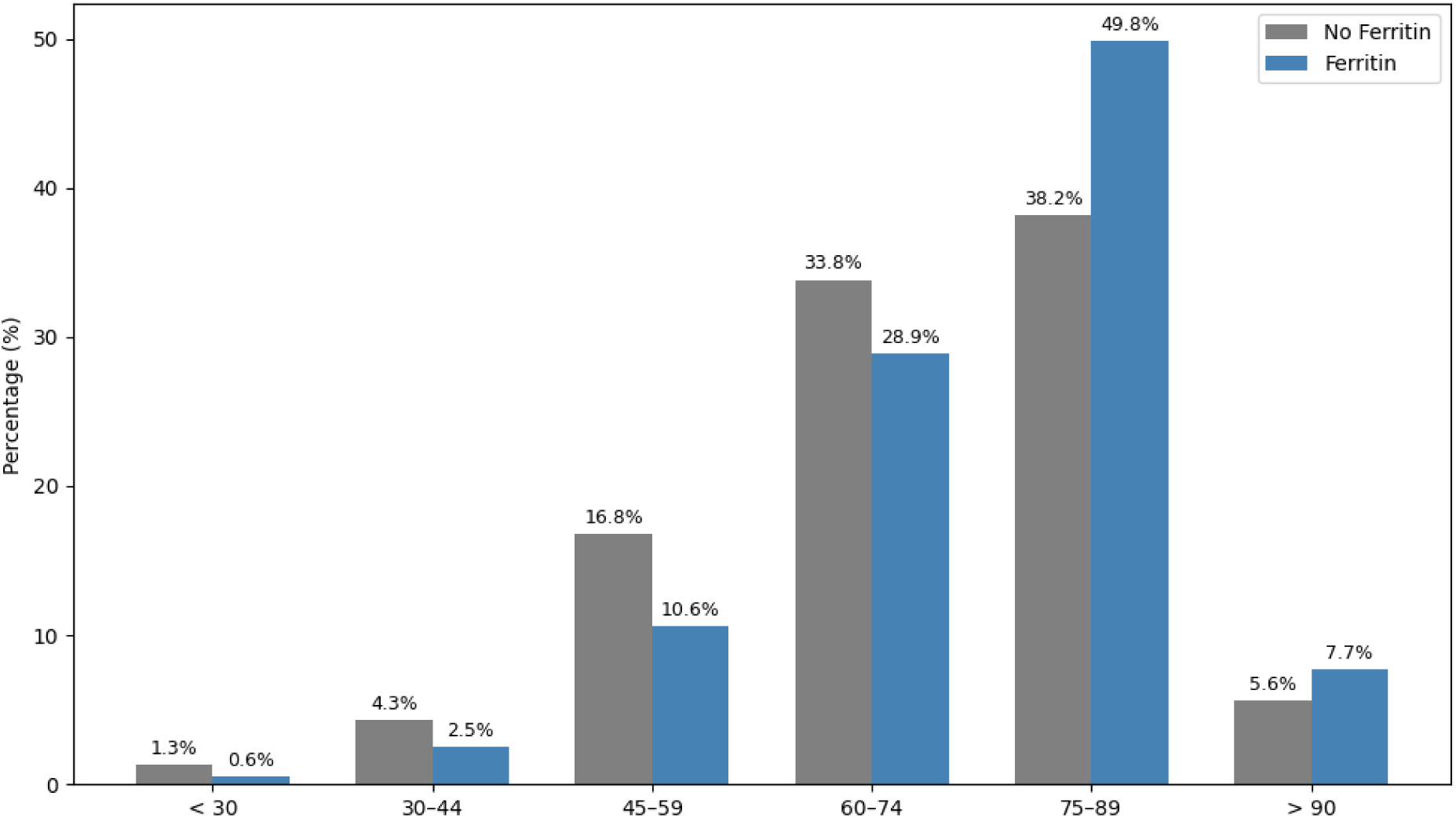
Proportion of people receiving a ferritin test by age categories.

**Table 2.**
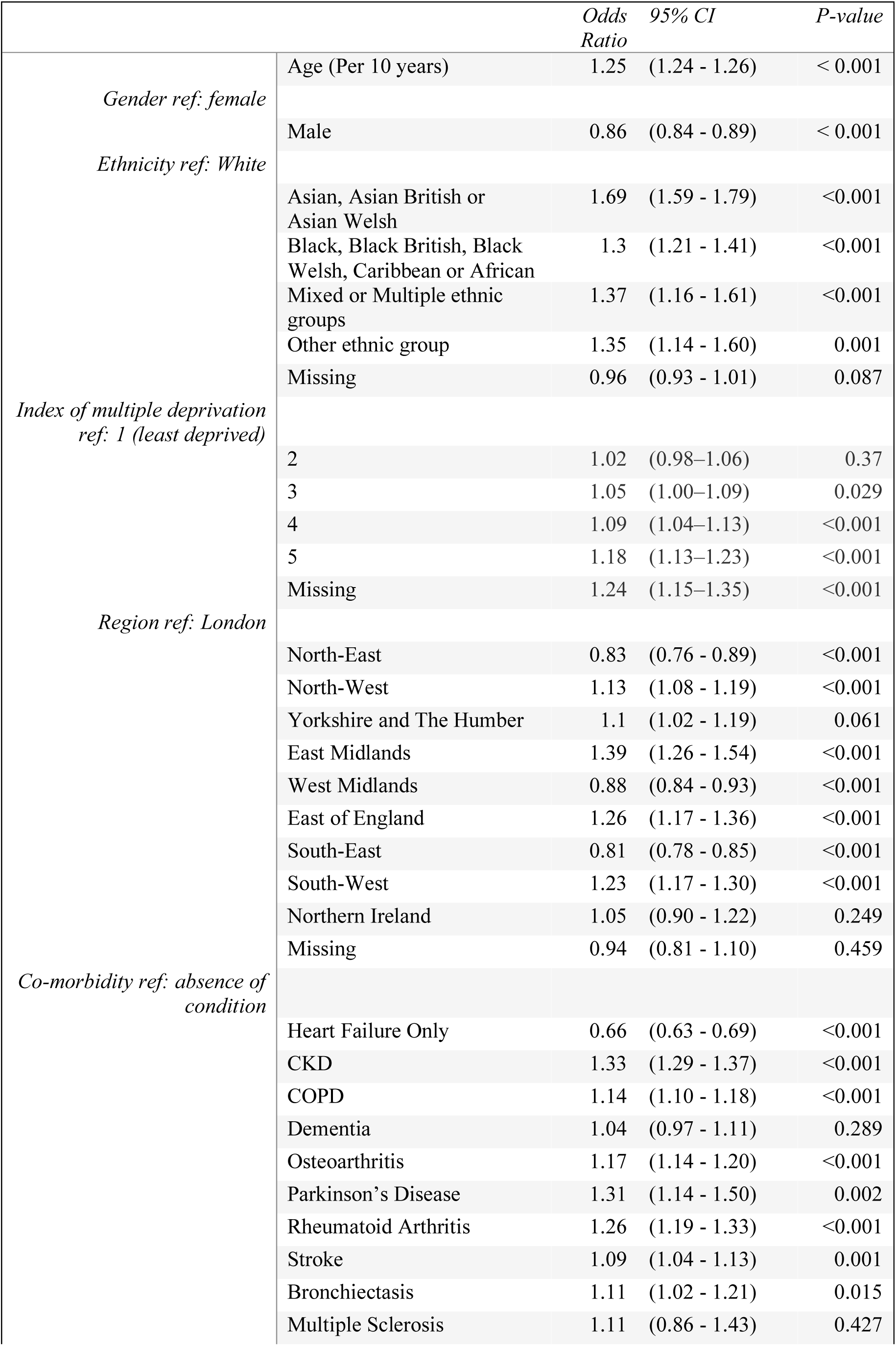
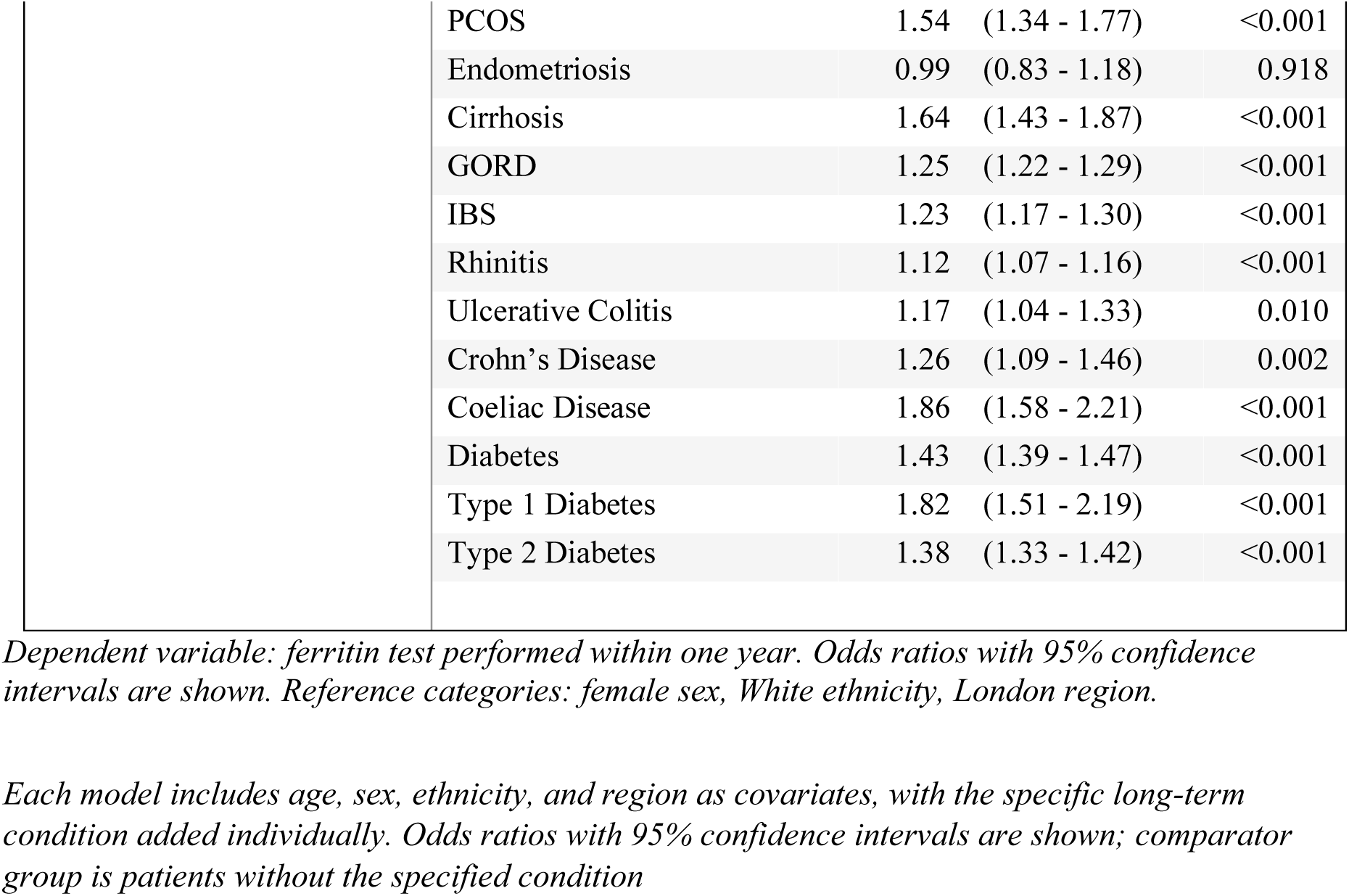
Multivariable logistic regression for predictors of ferritin testing within one year of incident heart failure diagnosis.

Substantial regional variation was observed in ferritin testing (Figure 3). The highest rates were in the East Midlands (41.6%), East of England (39.7%), and South-West (38.4%), while the North-East (28.5%), Northern Ireland (28.9%), and South-East (29.2%) had the lowest. Increasing deprivation was also associated with an increased odds of ferritin testing (OR per 1 quintile increase 1.04, 95%CI: 1.03-1.05). People in IMD 5 (most deprived) had an 18% higher odds of ferritin testing than IMD 1 (least deprived) (OR 1.18, 95%CI: 1.13-1.23). A greater proportion of people in IMD 5 also had MLTC; for example 15.0% (n=2,975) had four or more compared to 10.7% (n=2,291) in IMD 1. Overall, patients who underwent ferritin testing had a higher median number of long-term conditions (2 [IQR 1–3]) compared with those untested (1 [IQR 1–3]), a difference that was statistically significant (Mann–Whitney U test, p < 0.001). When the number of conditions was modelled categorically, the odds of ferritin testing rose progressively with additional comorbidities: from an OR of 1.25 (95% CI: 1.20–1.30) with one additional condition to 2.50 (95% CI: 1.25–4.87) with eight.

**Figure 3.**
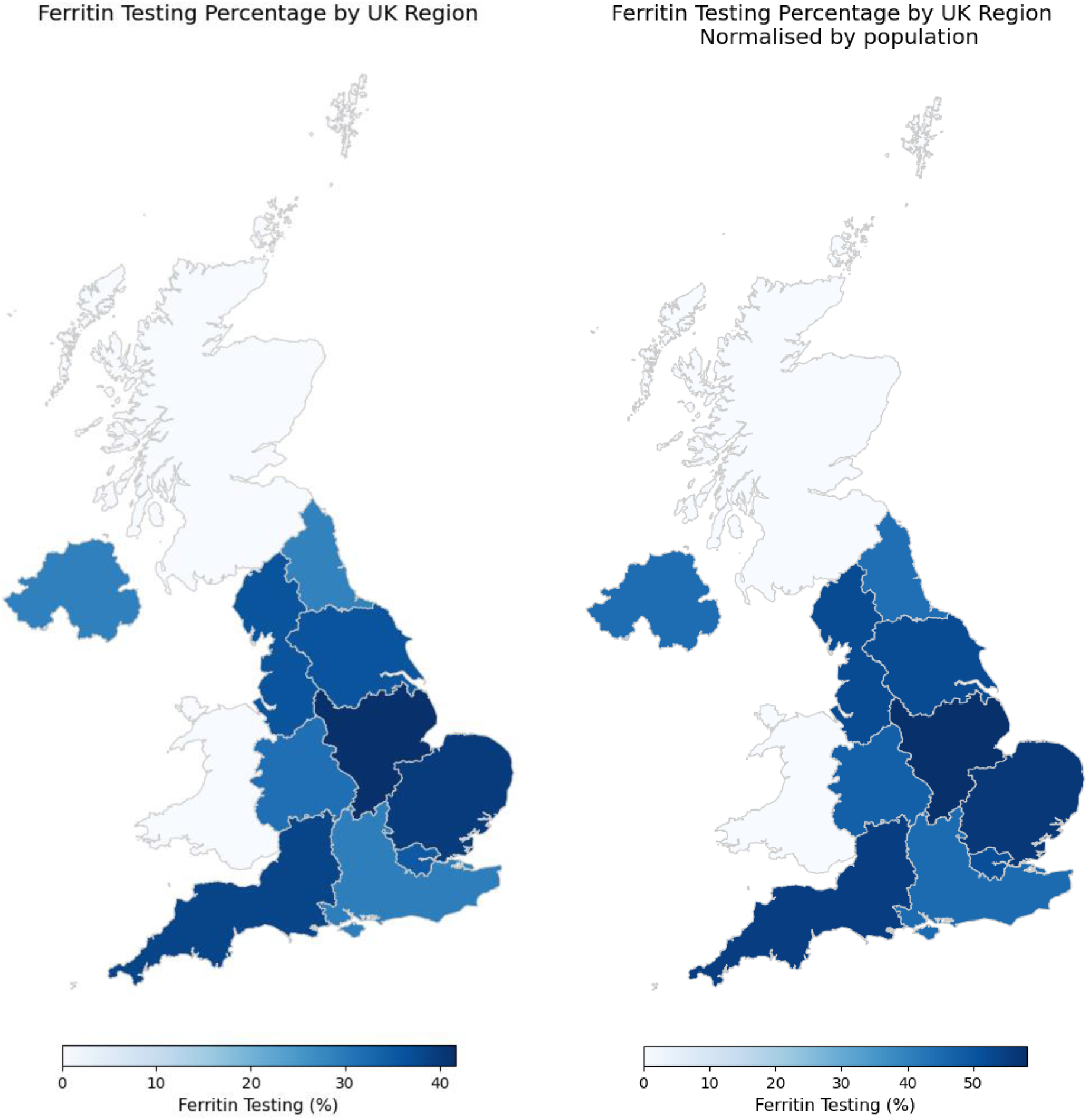
Regional variation in ferritin testing within the year following heart failure diagnosis. Left: Crude populations of patients tested by UK region. Right: Populations normalised by regional population

The odds of receiving a ferritin test were significantly higher based on the presence of all 19 pre-specified co-morbidities except for dementia, endometriosis and multiple sclerosis. The strongest associations were observed in individuals with coeliac disease (OR 1.86, 95%CI: 1.58-2.21), followed by those with type 1 diabetes (OR 1.82, 95%CI: 1.51-2.19) and cirrhosis (OR 1.64, 95%CI: 1.43-1.87) (Table 2). Patients with only heart failure and no other long-term conditions had the lowest likelihood of testing, with just 23.2% undergoing ferritin assessment within one year of diagnosis (OR 0.66, 95% CI: 0.63–0.69). Findings were consistent across complete case (Supplementary Table) and missing-indicator models, with no material change in the direction or strength of associations, indicating robustness to missing data.

### Ferritin monitoring

Only 13,309 (37.3%) of the 35,688 patients who had an initial ferritin test had a repeat measurement in the following year. Increased odds of repeat testing was associated with older age (OR 1.10 per 10-year increase in age, 95% CI: 1.08–1.12), Asian ethnicity (OR 1.25, 95%CI: 1.14-1.37) and female sex (OR for male sex 0.95, 95% CI: 0.91–1.00) (Table 3). Regional variation was again evident with the odds of repeat testing highest in the East Midlands (OR 1.36, 95% CI: 1.16–1.59) and South-West (OR 1.19, 95% CI: 1.09–1.29).

**Table 3.**
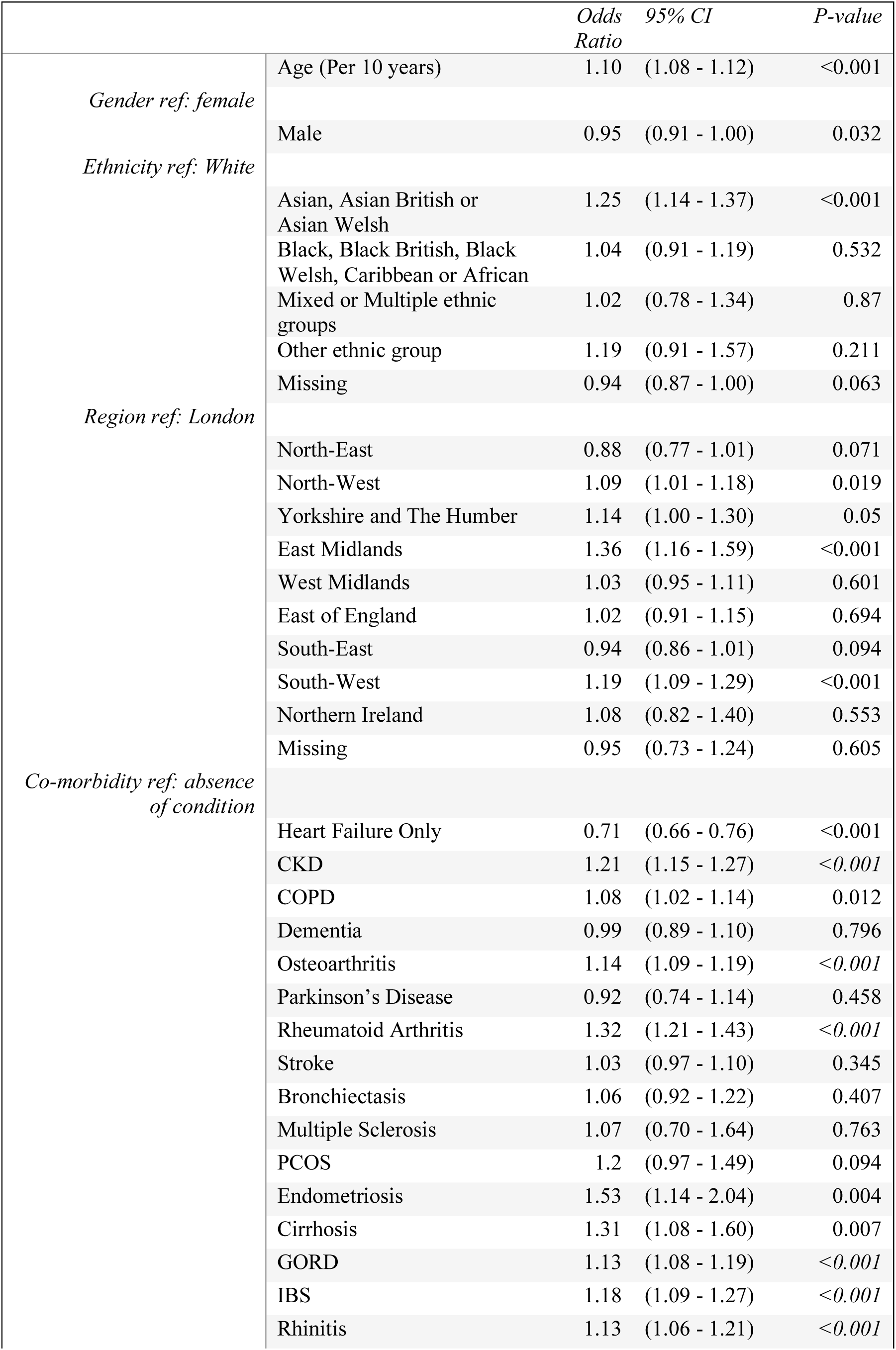
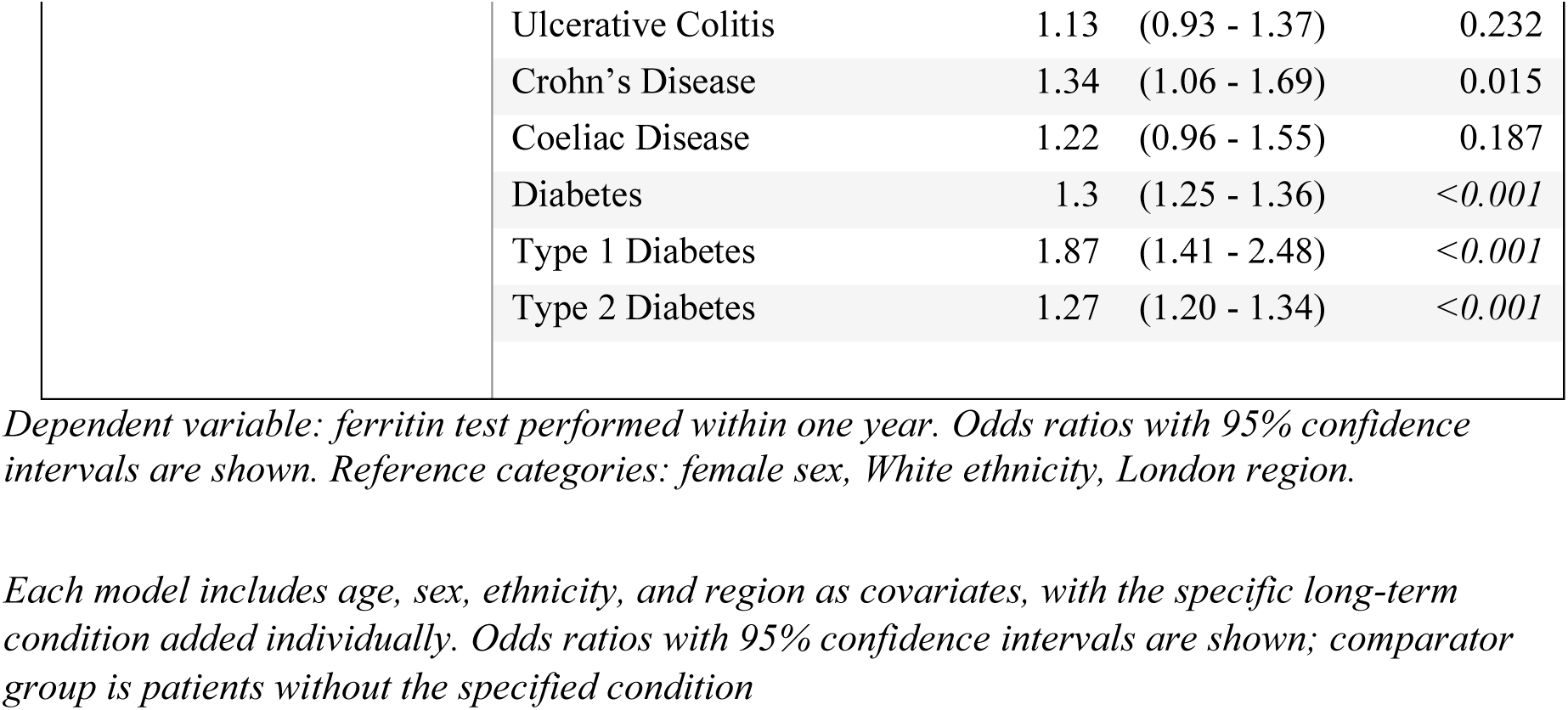
Multivariable logistic regression for predictors of <u>repeat</u> ferritin testing within one year of incident heart failure diagnosis.

Several long-term conditions were independently associated with increased odds of repeat ferritin testing (Table 3). The strongest associations were again observed for type 1 diabetes (OR 1.87, 95% CI: 1.41–2.48), coeliac disease (OR 1.34, 95% CI: 1.06–1.69) and cirrhosis (OR 1.31, 95% CI: 1.08–1.60), though in contrast to initial screening, the odds were also higher among people with endometriosis (OR 1.53, 95% CI: 1.14–2.04). Other gastrointestinal and inflammatory conditions, including Crohn’s disease, irritable bowel syndrome, and GORD, were also significantly associated with repeat testing. As with initial ferritin assessment, patients with heart failure as their only diagnosis remained significantly less likely to undergo repeat testing (OR 0.71, 95% CI: 0.66–0.76, p < 0.001). These findings were consistent in the complete case analysis (Supplementary table).

## Discussion

Within this large, retrospective primary care cohort we found that only a third of patients had a ferritin test within a year of an incident heart failure diagnosis, despite recommendations in international guidelines. Of those tested, only 13,309 (37.3%) had the ferritin level repeated. Younger people, males, people of White ethnicity and people who had heart failure without co-morbidities were less likely to be tested. Coeliac disease, cirrhosis and type 1 diabetes were associated with a higher odds of ferritin testing. Although a higher proportion of people in the least deprived socio-economic group were diagnosed with heart failure, a higher proportion in the most deprived quintile received a ferritin test, which may reflect an association between deprivation and increasing number of MLTCs. We also found there were regional variations in testing with the odds lowest for testing in the South-East and North-East and highest in the East of England and East Midlands. A similar pattern was seen for both initial ferritin testing and monitoring over time.

### Strengths and limitations

To our knowledge, this is the first study to report on ferritin testing among people with incident heart failure in primary care. A strength of our analysis is access to a large dataset, providing ‘real-world’ information in UK. Our analysis relies on the accuracy of primary care codes, which may not always be complete. For example, ferritin tests conducted in secondary care would not be captured unless manually transcribed to the primary care record so prevalence of testing could be under-estimated. However, it has been reported that at least half of people with chronic heart failure are exclusively managed in primary care, so it is likely that the low proportion of people undergoing ferritin testing is accurate.^19^ Most individuals did not have a code that categorised heart failure by left ventricular ejection fraction (LVEF). Iron replacement has particularly been tested as a treatment for HFrEF and it may be that some clinicians would base the decision as to whether to undertake ferritin testing based on the LVEF category but we were not able to report on this.

We focused on ferritin testing, but NICE guidelines recommend this is done alongside haemoglobin concentration and other biomarkers such as transferrin saturation (TSAT). Reporting on these tests might provide additional information around compliance with guidelines but does not impact on the primary results of our study. We did not capture the reason why patients had a ferritin test done, nor whether they had symptoms prior to testing. Many patients may have been having the ferritin tested for a reason other than heart failure, as although predominately as part of investigation of anaemia. Although a broader analysis may have been helpful in contextualising our results, this was beyond the scope of the current project.

Clinicians may also consider a patient’s overall health status and prognosis when considering ferritin testing. For some patients with advanced heart failure the focus of care may have been palliative or symptom relief leading to less testing. However, we excluded people who died within a year of their study index date to exclude those with a particularly poor prognosis.

### Comparison with existing literature

Previous primary care research has demonstrated that ferritin is a commonly requested test in primary care, irrespective of the presence of heart failure, with a recent Swiss study reporting 26.6% of individuals in 2018 received a test.^20^ This suggests that the presence of heart failure is only having a limited impact on testing and that many patients who received a ferritin test may have had this done anyway for another reason. There are known inequalities within in heart failure care in the UK, for example, older people and women experience longer delay from symptom presentation to heart failure diagnosis.^21^ However, we identified the inverse with ferritin testing. We found that older people were more likely to have a ferritin test, perhaps because the prevalence of anaemia increases with age.^22^ Older patients are also more likely to have increased number of long-term conditions, including CKD, for which testing of ferritin is more routine.^23^ Importantly, our results highlight young patients as a cohort at particular risk of under-testing with ferritin and so missed diagnoses.

### Implications for practice and research

Ferritin is an easily accessible test in primary care and iron replacement could improve quality of life. Yet, we found that only a third of patients with incident heart failure were screened as recommended for iron deficiency within the first year of diagnosis.^11^ ^14^ This represents a substantial shortfall in guideline-concordant care and highlights a persistent implementation gap within UK primary care. Reasons for non-testing need to be explored in further research, which may include low awareness of the guidance around ferritin testing, competing priorities in multimorbid patients, or uncertainty regarding the clinical significance of subclinical iron deficiency. Increased testing and earlier identification of iron deficiency could improve clinical outcomes and reduce healthcare costs associated with recurrent hospitalisation or functional decline.

## Acknowledgements

This study is based on data from the Clinical Practice Research Datalink (CPRD) obtained under licence from the UK Medicines and Healthcare products Regulatory Agency. The data is provided by patients and collected by the NHS as part of their care and support. The interpretation and conclusions contained in this study are those of the author alone. This work was carried out under approved study protocol 22_001873. This project is supported (in part) by the UK NIHR Blood and Transplant Research Unit in Data Driven Transfusion Practice (NIHR203334).

## Data availability

Requests to access data from CPRD are reviewed via the CPRD Research Data Governance process. Further information is available at: https://www.cprd.com/access-data.

